# The Gut Resistome during Hematopoietic Stem Cell Transplantation in Children

**DOI:** 10.1101/2022.07.07.22277185

**Authors:** Sarah M. Heston, Rebecca R. Young, Kirsten Jenkins, Paul L. Martin, Andre Stokhuyzen, Doyle V. Ward, Shakti K. Bhattarai, Vanni Bucci, Mehreen Arshad, Nelson J. Chao, Patrick C. Seed, Matthew S. Kelly

## Abstract

**Background:** Children undergoing hematopoietic stem cell transplantation (HCT) are at high risk of acquiring antibiotic-resistant bacteria. Few prior studies examined antibiotic resistance genes (ARGs) within the gut metagenomes of children undergoing HCT.

**Methods:** We conducted a longitudinal study of children (age <18 years) undergoing HCT at a single institution. We performed shotgun metagenomic sequencing of fecal samples collected between days -30 and +100 relative to HCT. We evaluated the effects of aerobic (cefepime, vancomycin, fluoroquinolones, aminoglycosides, macrolides, and trimethoprim-sulfamethoxazole) and anaerobic (piperacillin-tazobactam, carbapenems, metronidazole, and clindamycin) antibiotic exposures on the diversity and composition of the gut microbiome and resistome.

**Findings:** Using metagenomic data from 693 fecal samples collected from 80 children, we identified 350 unique ARGs. The most frequent ARGs identified encode resistance to tetracycline (n=91), beta-lactams (n=80), and fluoroquinolones (n=76). Both aerobic and anaerobic antibiotic exposures were associated with a decrease in the number of bacterial species (aerobic, β=0.72, 95% CI: 0.66, 0.79; anaerobic, β=0.68, 95% CI: 0.61, 0.76) and the number of unique ARGs (aerobic, β=0.83, 95% CI: 0.76, 0.91; anaerobic, β=0.84, 95% CI: 0.76, 0.93) within the gut metagenome. However, only anaerobic antibiotics were associated with an increase in the number of newly acquired ARGs (29%, 95% CI: 10%, 52%) and the abundance of ARGs (95%, 95% CI: 59%, 138%) in the gut resistome. Specific antibiotic exposures were associated with distinct changes in the number and abundance of resistance genes for individual antibiotic classes.

**Interpretation:** The gut metagenome and resistome of children are highly dynamic throughout HCT, driven largely by antibiotic exposures. Compared to antibiotics without anaerobic activity, anaerobic antibiotics were associated with increased microbiome instability and expansion of the gut resistome.

**Funding:** Antibacterial Resistance Leadership Group, National Institutes of Health, Duke Children’s Health & Discovery Initiative, Children’s Miracle Network Hospitals

## Introduction

Antibiotic resistance is one of the most serious global public health threats. In the United States alone, more than 2.8 million antibiotic-resistant infections occur each year, resulting in an estimated $4.6 billion in healthcare expenditures and more than 35,000 deaths.^1,2^ Despite the World Health Organization’s Global Action Plan to combat antimicrobial resistance, the number of infections and deaths from antimicrobial-resistant organisms continues to rise.^3^ Currently, 18 antimicrobial-resistant organisms are considered national threats by the United States Centers for Disease Control and Prevention, and there has been a 50% increase in the number of infections caused by extended-spectrum beta lactamase (ESBL)-producing Enterobacteriaceae alone since 2013.^1^ A number of factors contribute to growing antibiotic resistance, but use of antibiotics is the major driving force, thereby making hospitals a setting for the spread of antibiotic resistance among patients.^1,4^

Patients undergoing hematopoietic stem cell transplantation (HCT) are at high risk of acquiring antibiotic-resistant organisms due to their prolonged hospitalizations, impaired immunity, and frequent receipt of broad-spectrum antibiotics. Colonization by multidrug-resistant bacteria is observed in more than 50% of adult HCT recipients and has been associated with a higher incidence of non-relapse-related mortality.^5^ Moreover, HCT recipients are at high risk for antibiotic-resistant infections arising from the gut because of antibiotic-induced changes in the gut microbiome, impaired host immunity, and mucosal barrier injury from chemotherapy.^6,7^ Overgrowth of potential pathogens that colonize the gut increases the risk of an antibiotic-resistant bloodstream infection.^7,8^ In addition, antibiotics with an anaerobic spectrum of activity are increasingly recognized to disrupt the gut microbiome and are associated with poor outcomes after HCT including a higher risk of graft-versus-host disease (GVHD).^9^ Historically, studies of HCT recipients identified colonization by antibiotic-resistant bacteria using culture-or PCR-based methods. These methods are limited by focusing only on cultivable bacteria or on a small number of specific resistance genes. With the increasing accessibility of shotgun metagenomic sequencing, we are now able to simultaneously detect the full complement of antibiotic resistance genes (ARGs) within a sample, collectively referred to as the “resistome.”

In this study, we used shotgun metagenomic sequencing of 693 fecal samples from 80 children and adolescents undergoing HCT. We performed longitudinal analyses to evaluate the impact of aerobic and anaerobic antibiotics on gut microbiome composition and the gut resistome during HCT. In addition, we evaluated associations between specific antibiotic exposures and the number and abundances of resistance genes to specific antibiotic classes to describe the effect of specific antibiotic exposures on the gut resistome.

## Methods

### Study Design and Population

We conducted a prospective cohort study of children and adolescents (<18 years of age) who underwent their first HCT through the Duke University Pediatric Transplant and Cellular Therapy Program between October 2015 and February 2018. Subjects were prospectively enrolled during the pretransplantation evaluation, and fecal samples were collected from subjects on as many days as possible from enrollment through 100 days after HCT. Daily clinical data, including information on receipt of antibiotics, were collected from the electronic medical record. We obtained informed consent from participants’ legal guardians prior to enrollment, and the study protocol was approved by the Duke University Health System Institutional Review Board.

### Transplant Practices

Most children received trimethoprim-sulfamethoxazole (TMP-SMX) for *Pneumocystis jirovecii* prophylaxis from the start of the preparatory chemotherapy conditioning regimen through two days before HCT. Thereafter, children received monthly inhaled or IV pentamidine starting 30 days after HCT. After HCT, broad-spectrum antibiotics were initiated at onset of fever or concern for infection, but routine antibacterial prophylaxis was not used. Cefepime was the first-line antibiotic for subjects experiencing fever and neutropenia; vancomycin or antibiotics with an anaerobic spectrum of activity (e.g., piperacillin-tazobactam) were also frequently used if there was concern for a specific infectious source. Antibiotic selection and duration were at the discretion of the clinical provider.

### Metagenomic Sequencing and Data Processing

Approximately weekly fecal samples from each subject underwent shotgun metagenomic sequencing. DNA was extracted from fecal samples using PowerSoil Pro Kits (Qiagen, Germantown, MD). DNA sequencing libraries were constructed using Nextera XT DNA Library Prep Kits (Illumina, San Diego, CA) and sequenced on NextSeq500 or NovaSeq6000 instruments (Illumina) as 150-bp paired-end reads. We trimmed reads using Trimmomatic (version 0.39) and removed host decontamination using Bowtie2 (version 2.3.5).^10,11^ Host-decontaminated reads were profiled for bacterial species relative abundances using MetaPhlAn2.^12^ Using the ShortBRED pipeline (version 0.9.4), we aligned sequencing reads to the Comprehensive Antibiotic Resistance Database (CARD; version 313).^13,14^ Reads mapped to antibiotic resistance genes were normalized by dividing the proportion of sequencing reads mapped to a gene sequence by the length of the gene sequence to determine the reads per kilobase per million (RPKM). We pruned samples with less than 500,000 quality-filtered metagenomic read-pairs.

### Statistical Analyses

We used MaAsLin2 to fit linear mixed effects models with subject as a random effect to evaluate associations between antibiotic exposures and clinical characteristics and the relative abundances of specific bacterial species.^15^ For these models, we considered species that had a minimum mean relative abundance of 1% and a sample prevalence of 5%; the false discovery rate was set to 0.1. We considered cefepime, vancomycin, fluoroquinolones, aminoglycosides, macrolides, and TMP-SMX to have aerobic spectra of activity. We considered piperacillin-tazobactam, carbapenems, metronidazole, and clindamycin to have spectra of activity that included anaerobes. A participant was considered exposed to an antibiotic if they received at least one dose of the antibiotic since the last sequenced sample. If a subject started antibiotics on the day of fecal sample collection, the antibiotic exposure was assigned to the next sequenced sample.

We assessed for the effects of time during HCT and antibiotic exposures on gut microbiome composition and the gut resistome using mixed effects models with day relative to HCT, aerobic antibiotic exposure, and anaerobic antibiotic exposure as fixed effects and subject as a random effect. Outcome measures for these models included the number of species and ARGs, the number of new species and ARGs acquired since the previous sample, microbiome and resistome instability measured by the Jaccard distance, and the abundance of ARGs.^16^ Models were adjusted for subject age, sex, underlying diagnosis, preparatory conditioning regimen, type of HCT (autologous or allogeneic), and sample sequencing depth. Models for the number of species and ARGs and the number of newly acquired species and ARGs were fit using a negative binomial distribution with the *glmmADMB* R package (version 0.8.3.3). Models for the Jaccard distance were fit using linear regression with the *lme4* R package (version 1.1-27.1).^17,18^ We performed similar analyses to evaluate associations between exposures to specific antibiotics and measures of the resistome using adjusted mixed effects models. We correlated measures of the resistome with corresponding measures of microbiome composition using repeated measures correlations implemented using the *rmcorr* R package (version 0.4.4).^19^ Finally, we considered the effect of individual antibiotics on the number and abundance of ARGs that confer resistance to specific classes of antibiotics using unadjusted mixed effect negative binomial models with individual antibiotic exposure as a fixed effect and subject as a random effect. We applied the Benjamini-Hochberg correction for multiple comparisons on the unadjusted models.^20^ All analyses were conducted using R statistical software (version 4.0.2).

## Results

### Patient and sample characteristics

Our cohort included 80 children with a median (interquartile range [IQR]) age of 5.1 (2.2, 12.8) years (**Table 1**). The most frequent indication for HCT was hematologic malignancy (42%), and most transplants were allogeneic (88%), with umbilical cord blood being the most common donor source (64%). Cefepime, vancomycin, and TMP-SMX were the most commonly received antibiotics (**Table 2**). With regard to antibiotics with an anaerobic spectrum of activity, 44 (55%) subjects received metronidazole, 26 (33%) received piperacillin-tazobactam, 15 (19%) received a carbapenem, and four (5%) received clindamycin. Analyses included metagenomic sequencing data from 693 fecal samples [median (IQR) of 8 (6, 12) samples per child]. Shotgun metagenomic sequencing generated a total of 5,624,857,872 high-quality, paired-end metagenomic reads with a median (IQR) sequencing depth of 7.45 (3.41, 12.02) million reads per sample.

**Table 1.**
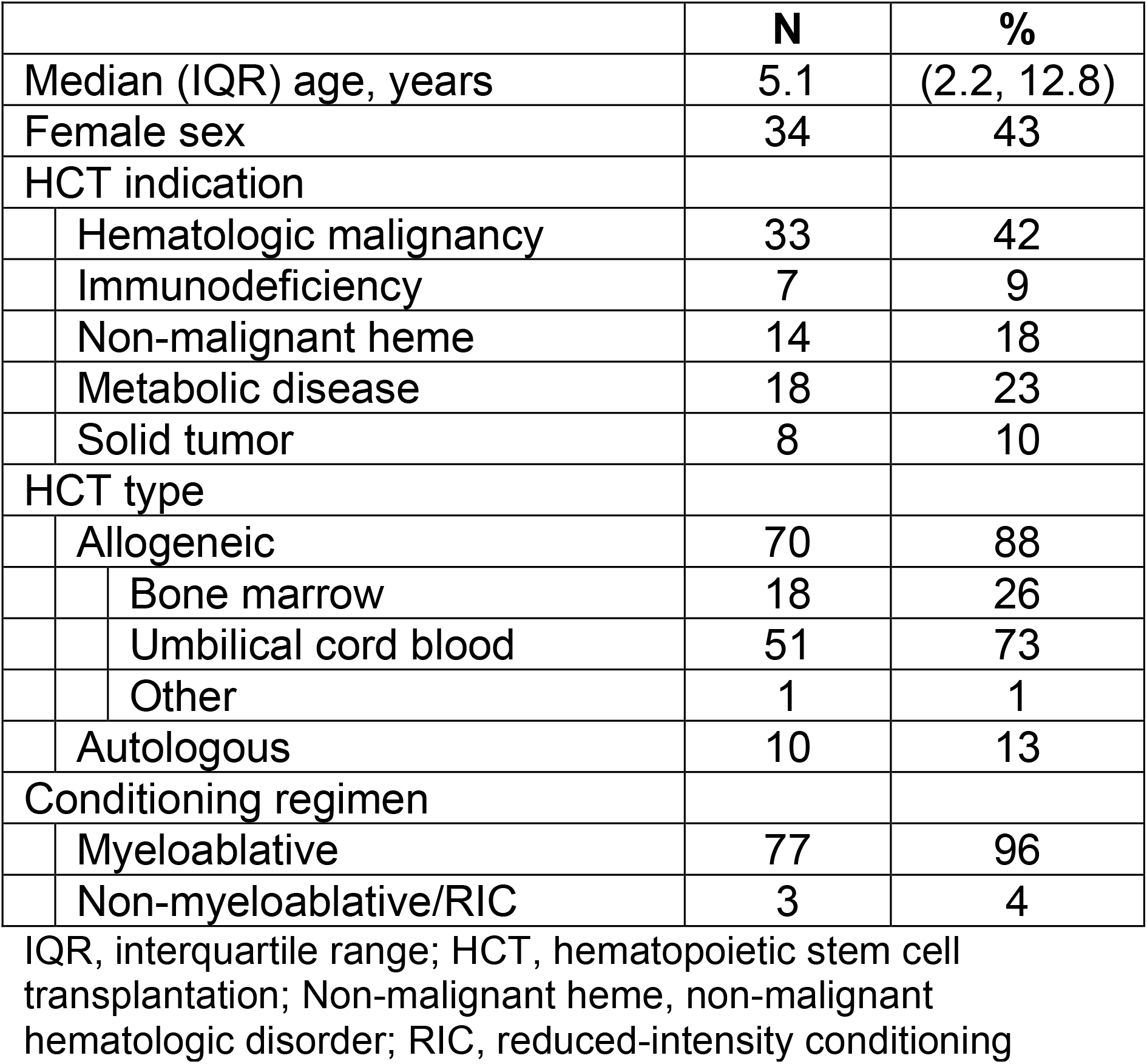
Characteristics of the study population of 80 children and adolescents undergoing HCT

|  | **N** | **%** |
| --- | --- | --- |
| Median (IQR) age, years | 5.1 | (2.2, 12.8) |
| Female sex | 34 | 43 |
| HCT indication |  |  |
| Hematologic malignancy | 33 | 42 |
| Immunodeficiency | 7 | 9 |
| Non-malignant heme | 14 | 18 |
| Metabolic disease | 18 | 23 |
| Solid tumor | 8 | 10 |
| HCT type |  |  |
| Allogeneic | 70 | 88 |
| Bone marrow | 18 | 26 |
| Umbilical cord blood | 51 | 73 |
| Other | 1 | 1 |
| Autologous | 10 | 13 |
| Conditioning regimen |  |  |
| Myeloablative | 77 | 96 |
| Non-myeloablative/RIC | 3 | 4 |

IQR, interquartile range; HCT, hematopoietic stem cell transplantation; Non-malignant heme, non-malignant hematologic disorder; RIC, reduced-intensity conditioning

**Table 2.** Prevalences of antibiotic exposures among the study population and fecal samples collected after antibiotic exposures

| **Antibiotic** | **Subject (N=80)** |  | **Samples (N=693)** |  |
| --- | --- | --- | --- | --- |
|  | **N** | **%** | **N** | **%** |
| Cefepime | 72 | 90 | 258 | 37 |
| Vancomycin | 73 | 91 | 176 | 25 |
| TMP-SMX | 71 | 89 | 162 | 23 |
| Metronidazole | 44 | 55 | 99 | 14 |
| Pip-Tazo | 26 | 33 | 66 | 10 |
| Macrolide | 19 | 24 | 36 | 5 |
| Other Beta-Lactam | 15 | 19 | 58 | 8 |
| Carbapenem | 15 | 19 | 27 | 4 |
| Aminoglycoside | 10 | 13 | 13 | 2 |
| Fluoroquinolone | 8 | 10 | 32 | 5 |
| Clindamycin | 4 | 5 | 11 | 2 |
| Tetracycline | 1 | 1 | 3 | 0 |

TMP-SMX, trimethoprim-sulfamethoxazole; Pip-Tazo, piperacillin-tazobactam

### Composition of the gut microbiome during HCT

We identified a total of 597 bacterial species with a median (IQR) of 28 (15, 50) unique bacterial species per sample. The most highly abundant bacterial species were *Escherichia coli* (mean relative abundance [MRA]=0.067), *Enterococcus faecalis* (MRA=0.060), *Bacteroides vulgatus* (MRA=0.056), *Enterococcus faecium* (MRA=0.039), and *Bacteroides stercoris* (MRA=0.033; **Figure 1**). The composition of the gut microbiome underwent substantial changes during the course of HCT; specifically, the relative abundances of several anaerobic species declined and the relative abundances of potential pathogens, including *Klebsiella pneumoniae, Enterococcus faecalis*, and several of the viridans group streptococci, increased during the course of HCT (**Supplementary Table 1**). The number of acquired bacterial species increased by 1% (95% confidence interval [CI]: 0.7%, 1.1%) per day after HCT (**Supplementary Table 2**). There were no significant changes in the total number of species or the instability of the gut microbiome with increasing time since HCT.

**Figure 1.**
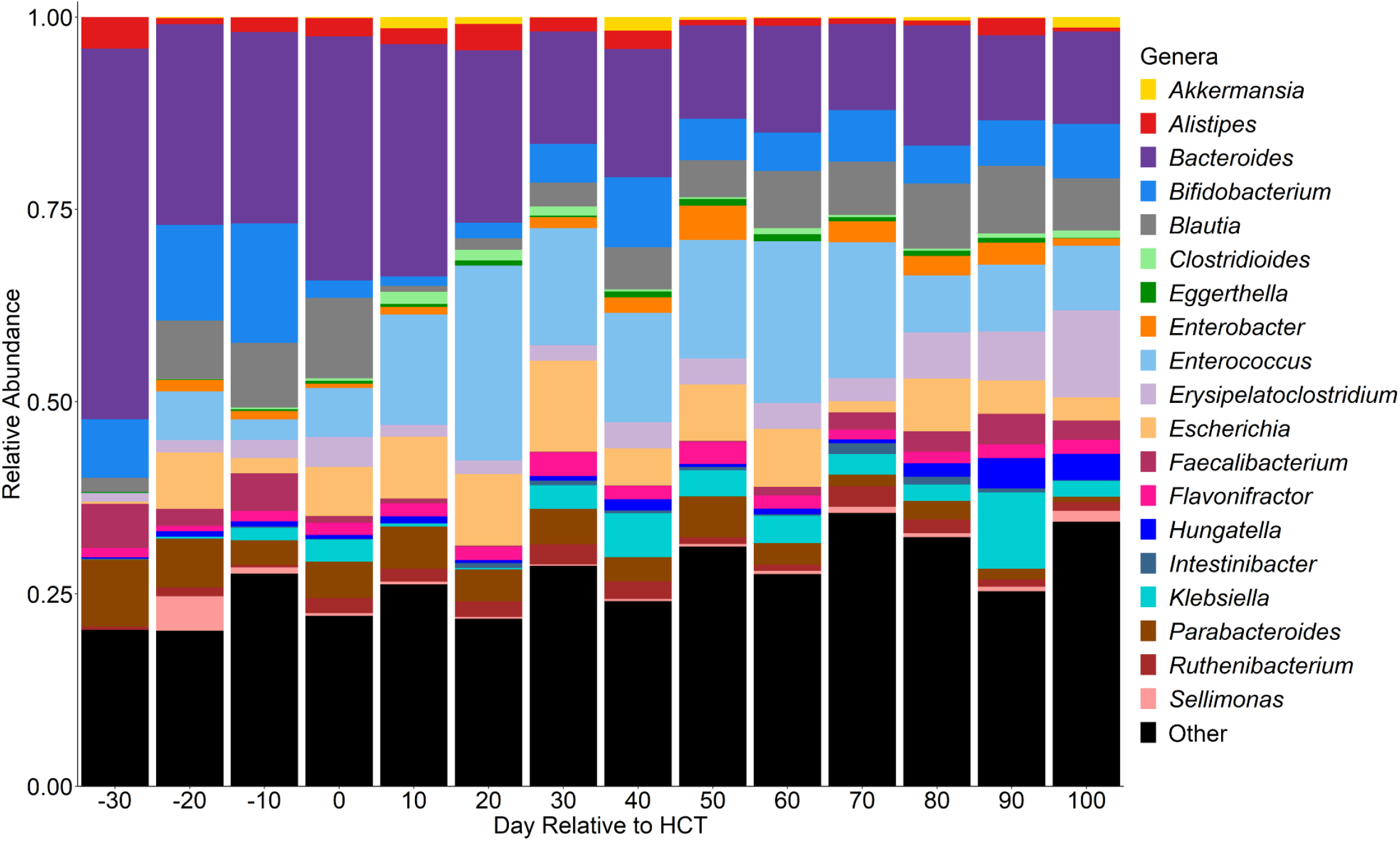
Relative abundances of highly abundant genera over time. Relative abundances of highly abundant genera fecal samples of children by day relative to hematopoietic stem cell transplantation.

Exposure to aerobic antibiotics was associated with a 28% (95% CI: 21%, 34%) decrease in the number of species and a 0.08 (95% CI: 0.04, 0.12) increase in instability. Anaerobic antibiotic exposures were similarly associated with a 32% (95% CI: 24%, 39%) decrease in the number of species and a 0.09 (0.05, 0.13) increase in instability of the gut microbiome composition (**Supplementary Table 2**). However, while aerobic antibiotics had no effect on the number of acquired species, there was a 19% (95% CI: 2%, 39%) increase in the number of acquired species associated with anaerobic antibiotics (**Supplementary Table 2**). Taken together, both aerobic and anaerobic antibiotic exposures were associated with a decrease in the number of bacterial species in the gut microbiome and an increase in instability of gut microbiome composition, but only anaerobic antibiotic exposures were associated with an increase in the acquisition of new species.

We next sought to evaluate associations between exposures to specific antibiotics and changes in the composition of the gut microbiome (**Figure 2**). Cefepime exposure was associated with decreases in the relative abundances of both anaerobic bacteria from the genera *Bifidobacterium, Blautia*, and *Clostridium*, as well as potential pathogens including *Escherichia coli, Klebsiella pneumoniae*, and viridans group streptococci. Piperacillin-tazobactam and metronidazole, both anaerobic antibiotics, were associated with decreases in the relative abundances of several anaerobic bacterial species. Piperacillin-tazobactam was also associated with an increase in the relative abundance of *Enterococcus faecium*, and metronidazole was associated with increases in the relative abundances of *E. faecalis* and *Enterococcus gallinarum*. Notably, fluoroquinolone exposure tended to be associated with increases in the relative abundances of several bacterial species from the genera *Bacteroides, Bifidobacterium, Blautia*, and *Clostridium* (**Supplementary Table 1**).

**Figure 2.**
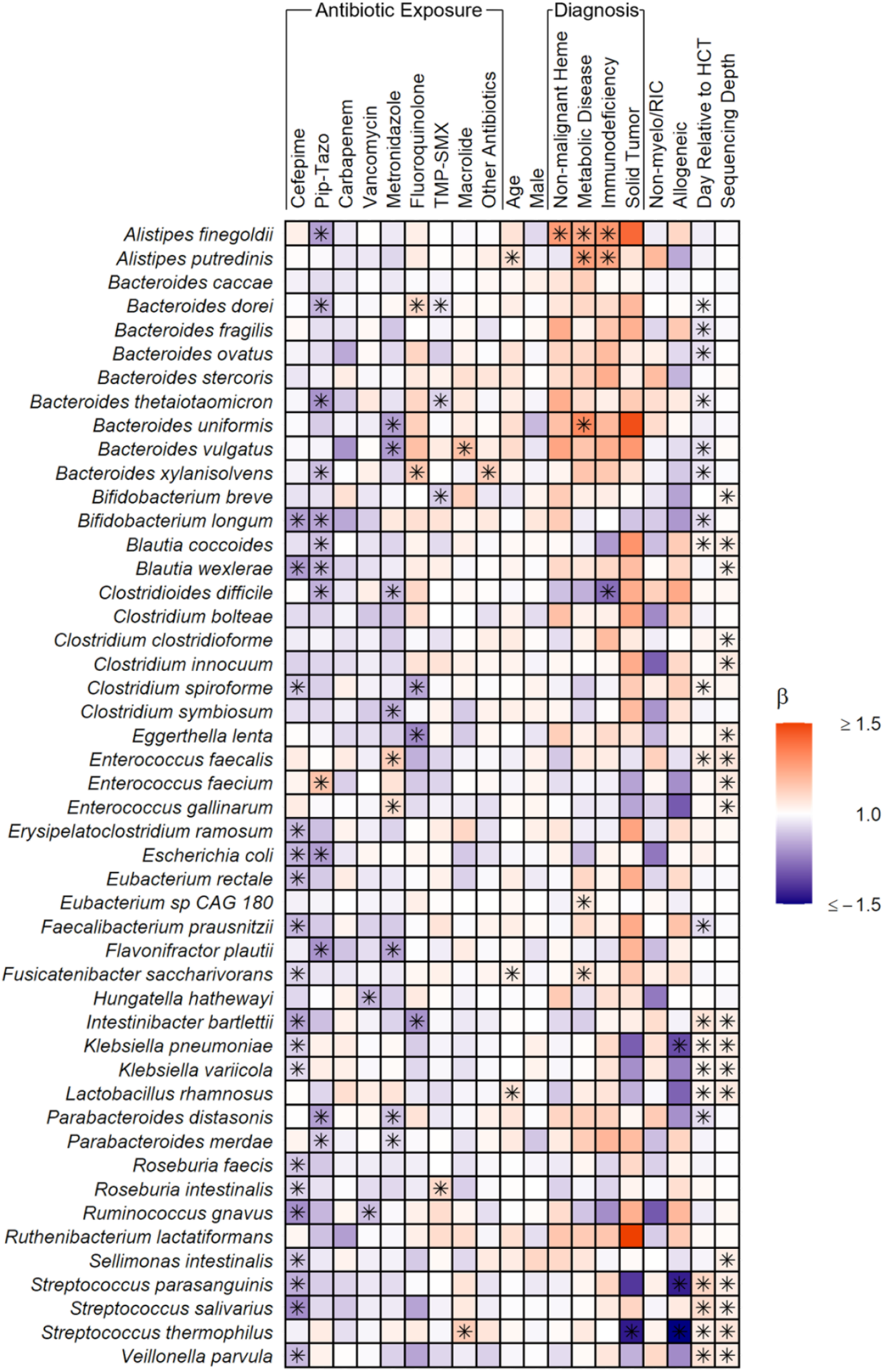
Heatmap depicting associations between antibiotic exposures and other clinical factors and gut microbiome composition. Associations were estimated from linear mixed effects models fit using MaAsLin2. We included bacterial species with a minimum mean relative abundance of 1% and a sample prevalence of 5%; the false discovery rate was set to 0.1. “Other” antibiotics includes exposures to tetracyclines, clindamycin, aminoglycosides, and non-cefepime beta-lactams. Betas indicate effect sizes and asterisks denote significant changes. Pip-Tazo, piperacillin-tazobactam; TMP-SMX, trimethoprim-sulfamethoxazole; Non-malignant heme, Non-malignant hematologic disorder; Non-myelo/RIC, non-myeloablative and reduced-intensity conditioning.

### The gut resistome during HCT

We detected a total of 350 unique ARGs with a median (IQR) of 28 (15, 56) ARGs per sample. The most common ARGs detected were ErmB, tetO, tetW, and dfrF, each of which were present in more than 60% of samples and can confer resistance to macrolides and lincosamides (e.g., clindamycin), tetracyclines, and trimethoprim, respectively. The most common ARG classes were tetracyclines, beta-lactams, and fluoroquinolones (**Table 3**).

**Table 3.** Antibiotic resistance genes identified in fecal samples that may confer resistance to clinically relevant antibiotics

| **Resistance Gene Class** | **Number of Genes Detected** | **Median (IQR) Number of ARGs per Sample** | **Median (IQR) ARG Abundance per Sample (RPKM)** |
| --- | --- | --- | --- |
| Tetracycline<sup>a</sup> | 94 | 9 (5, 15) | 328 (159, 623) |
| Beta-lactam<sup>b</sup> | 80 | 5 (1, 17) | 154 (19, 518) |
| Fluoroquinolone | 76 | 6 (1, 19) | 106 (5, 470) |
| Macrolide | 74 | 6 (4, 12) | 247 (107, 623) |
| Aminoglycoside | 67 | 4 (1, 8) | 101 (20, 373) |
| Phenicol | 64 | 3 (1, 6) | 48 (7, 202) |
| Carbapenem<sup>c</sup> | 36 | 1 (0, 3) | 8 (0, 81) |
| Lincosamide<sup>d</sup> | 31 | 3 (2, 5) | 124 (41, 254) |
| Diaminopyrimidine<sup>e</sup> | 30 | 2 (1, 3) | 42 (10, 128) |
| Glycopeptide<sup>f</sup> | 29 | 0 (0, 1) | 0 (0, 5) |
| Oxazolidinone<sup>g</sup> | 11 | 1 (0, 2) | 7 (0, 37) |
| Sulfonamide | 8 | 0 (0, 1) | 1 (0, 2) |
| Nitromidazole<sup>h</sup> | 1 | 0 (0, 1) | 0 (0, 35) |

IQR, interquartile range; RPKM, reads per kilobase million mapped reads
<sup>a</sup>Tetracycline includes tetracycline and glycylcycline (e.g., tigecycline) resistance genes
<sup>b</sup>Beta-lactam includes cephalosporin, cephamycin, penam, and monobactam resistance genes
<sup>c</sup>Carbapenem includes carbapenem and penem resistance genes
<sup>d</sup>Lincosamide includes clindamycin resistance genes
<sup>e</sup>Diaminopyrimidine includes trimethoprim resistance genes
<sup>f</sup>Glycopeptide includes vancomycin resistance genes
<sup>g</sup>Oxazolidinone includes linezolid resistance genes
<sup>h</sup>Nitromidazole includes metronidazole resistance genes

The gut resistome of children was also highly dynamic during the course of HCT. There was no significant change in the relative abundance of ARGs within the gut. However, there was a 0.2% (95% CI: 0.1%, 0.3%) increase in the number of ARGs and a 0.8% (95% CI: 0.6%, 1.0%) increase in the number of new ARGs with each day after HCT. Interestingly, the instability of the resistome decreased with each day after HCT (β=-0.001; 95% CI: -0.002, -0.000; **Figure 3; Supplementary Table 2**). While each year of increasing age was associated with a 5% (95% CI: 1%, 8%) decrease in gut resistome abundance and a decrease in resistome instability (β=-0.01; 95% CI: - 0.01, -0.00), other subject and transplant factors were not associated with measures of the resistome (**Supplementary Table 2**). Thus, when age and other clinical factors are held constant, new ARGs are acquired during HCT and the overall stability of the resistome increases with time.

**Figure 3.**
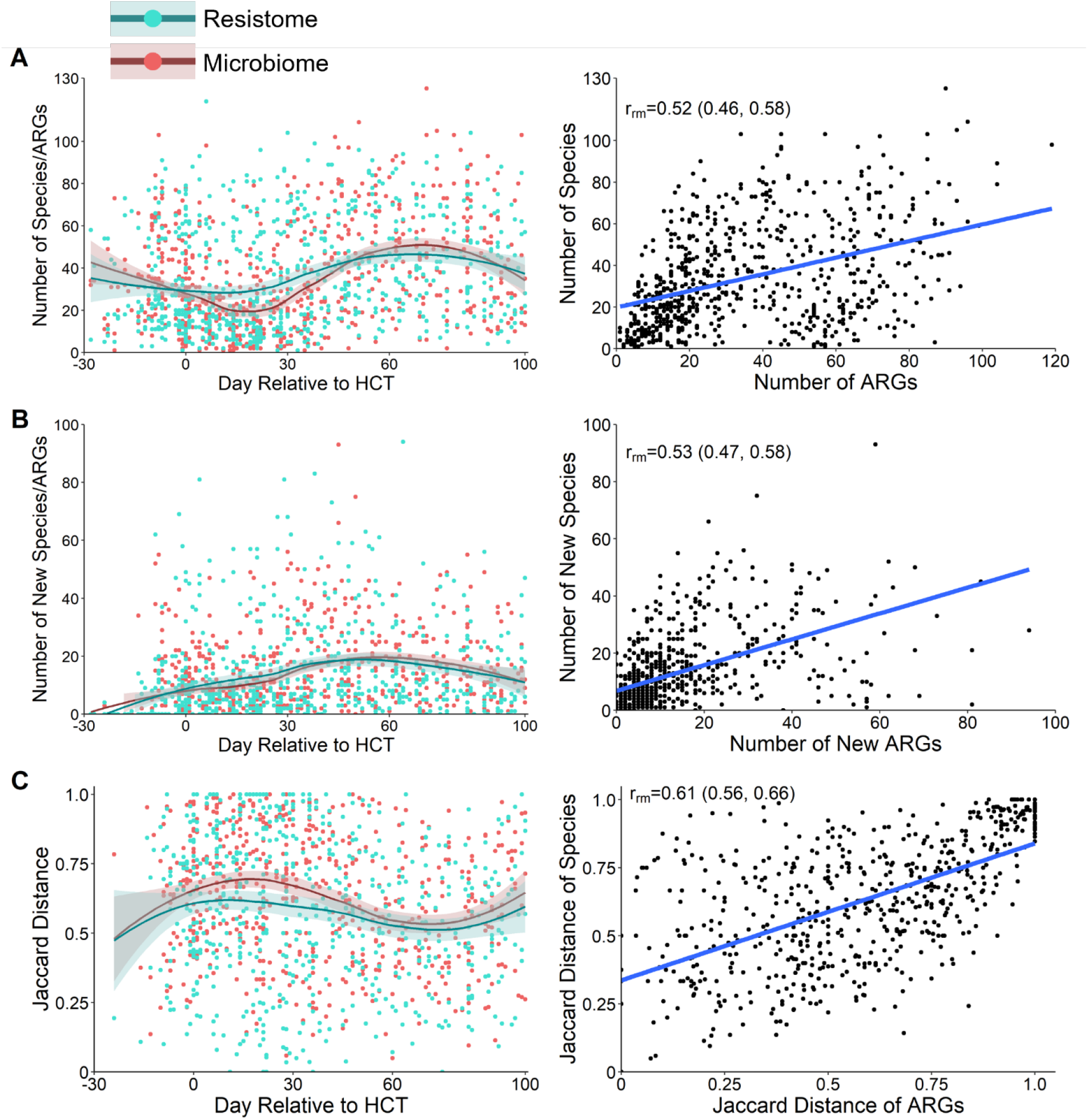
Changes to the gut resistome and microbiome over time. A. Number of ARGs from the resistome and number of bacterial species from the microbiome in fecal samples by the day relative to HCT; correlation of the number of bacterial species and number of ARGs for each sample. B. Number of new ARGs from the resistome and bacterial species from the microbiome by the day relative to HCT; correlation of the number of new bacterial species and ARGs since the previous sample. C. Jaccard distance in ARGs from the resistome and bacterial species from the microbiome by day relative to HCT; correlation of the Jaccard distances of the microbiome and resistome. The Jaccard distance is a measure of dissimilarity between two samples, with values closer to one representing increased instability.^16^ Points represent individual fecal samples; the smoothed line represents the median value over time, and shaded area represents the 95% confidence interval. Resistome is represented in blue, and the microbiome in red. ARG, antibiotic resistance gene; HCT, hematopoietic stem cell transplantation; r_rm_, repeated measures correlation coefficient.

Antibiotic exposures were a main driver of changes in the gut resistome in pediatric HCT recipients. Compared to no aerobic antibiotic exposure, aerobic antibiotics were associated with a 17% (95% CI: 9%, 24%) decrease in the number of ARGs but not with a change in the number of acquired ARGs, the abundance of ARGs, or the instability of ARGs (**Supplementary Table 2**; **Figure 4**). Conversely, anaerobic antibiotic exposure led to a 16% (95% CI: 7%, 24%) decrease in the number of ARGs, a 29% (95% CI: 10%, 52%) increase in the number of new ARGs, a 95% (95% CI: 59%, 138%) increase in the abundance of ARGs, and a 0.07 (95% CI: 0.03, 0.12) increase in the instability of ARGs compared to no anaerobic antibiotic exposure (**Figure 4**). When considered individually, there were antibiotic-specific effects on measures of the resistome (**Figure 4**; **Supplementary Table 3**). Thus, while both aerobic and anaerobic antibiotics led to a decrease in the number of ARGs, only anaerobic antibiotic exposures were associated with more acquired ARGs and higher ARG abundances.

**Figure 4.**
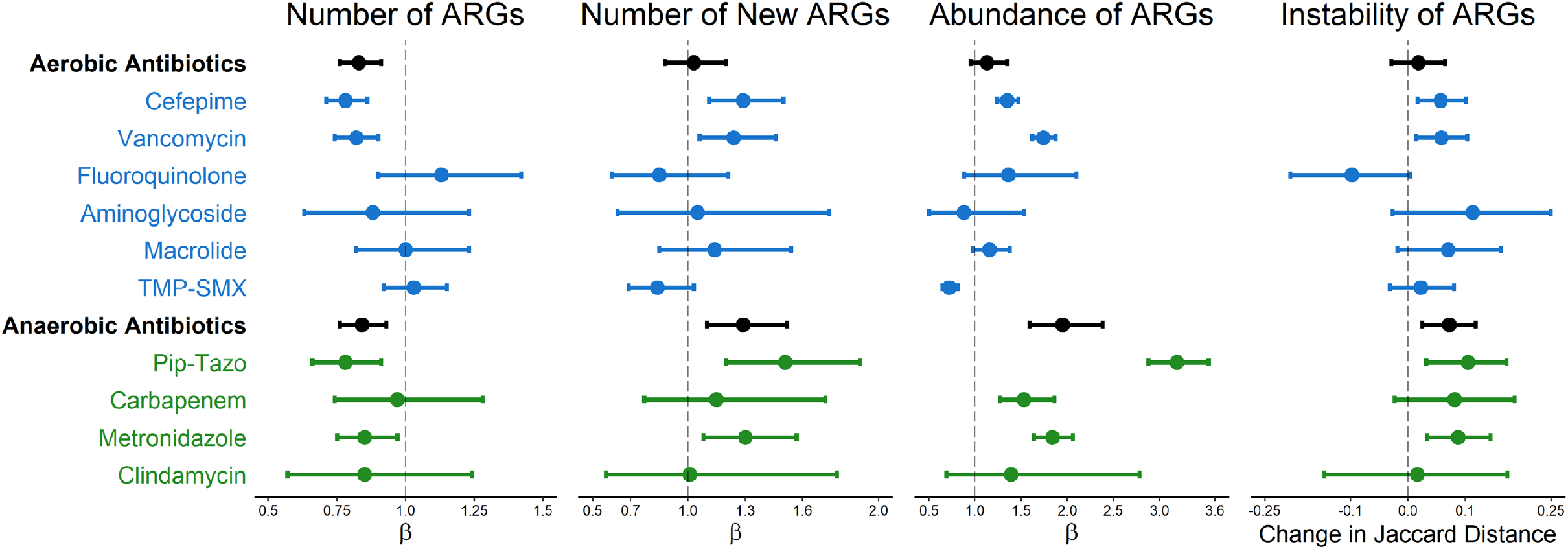
Effect of aerobic and anaerobic antibiotics on measures of the resistome. The effect of aerobic and anaerobic antibiotics for each measure of the resistome: the number of ARGs per sample, the number of new ARGs per sample, the abundance of ARGs, and the instability of the ARGs as measured by the Jaccard distance. The Jaccard distance is a measure of dissimilarity between two samples, with values closer to one representing increased instability.^16^ Points represent estimates, and error bars denote the 95% confidence interval. ARG, antibiotic resistance gene; TMP-SMX, trimethoprim-sulfamethoxazole; Pip-Tazo, piperacillin-tazobactam.

We next evaluated for correlations between measures of gut microbiome composition and the gut resistome using repeated measures correlation. There were moderate correlations between the number of ARGs and bacterial species (r_rm_=0.52; 95% CI: 0.46, 0.58), the number of acquired ARGs and bacterial species (r_rm_=0.53; 95% CI: 0.47, 0.58), and the stability of the gut microbiome and resistome (r_rm_=0.61; 95% CI: 0.56, 0.66; **Figure 3**). Taken together, these data suggest that the richness of the gut microbiome composition if a major driver of the gut resistome, and losses of gut microbial richness – as occurs with antibiotic exposure – in turn reduces the richness of the gut resistome.

### Antibiotic Exposures Affect Same and Different ARG Classes

Finally, we determined the effect of specific antibiotic exposures on the number and abundance of ARGs of the same antibiotic class and other ARG classes (**Figure 5; Supplementary Tables 4 and 5**). Cefepime was associated with decreases in the number and abundance of ARGs for multiple ARG classes, including ARGs that confer resistance to beta-lactams, carbapenems, and fluoroquinolones. Piperacillin-tazobactam and metronidazole, both anaerobic antibiotics, led to a decrease in the number of ARGs from many antibiotic classes but with increases in the relative abundances of some classes of ARGs, especially glycopeptide ARGs that confer resistance to vancomycin. Fluoroquinolone exposure tended to be associated with an increased number of ARGs and an increased relative abundance of ARGs for several antibiotic classes.

**Figure 5.**
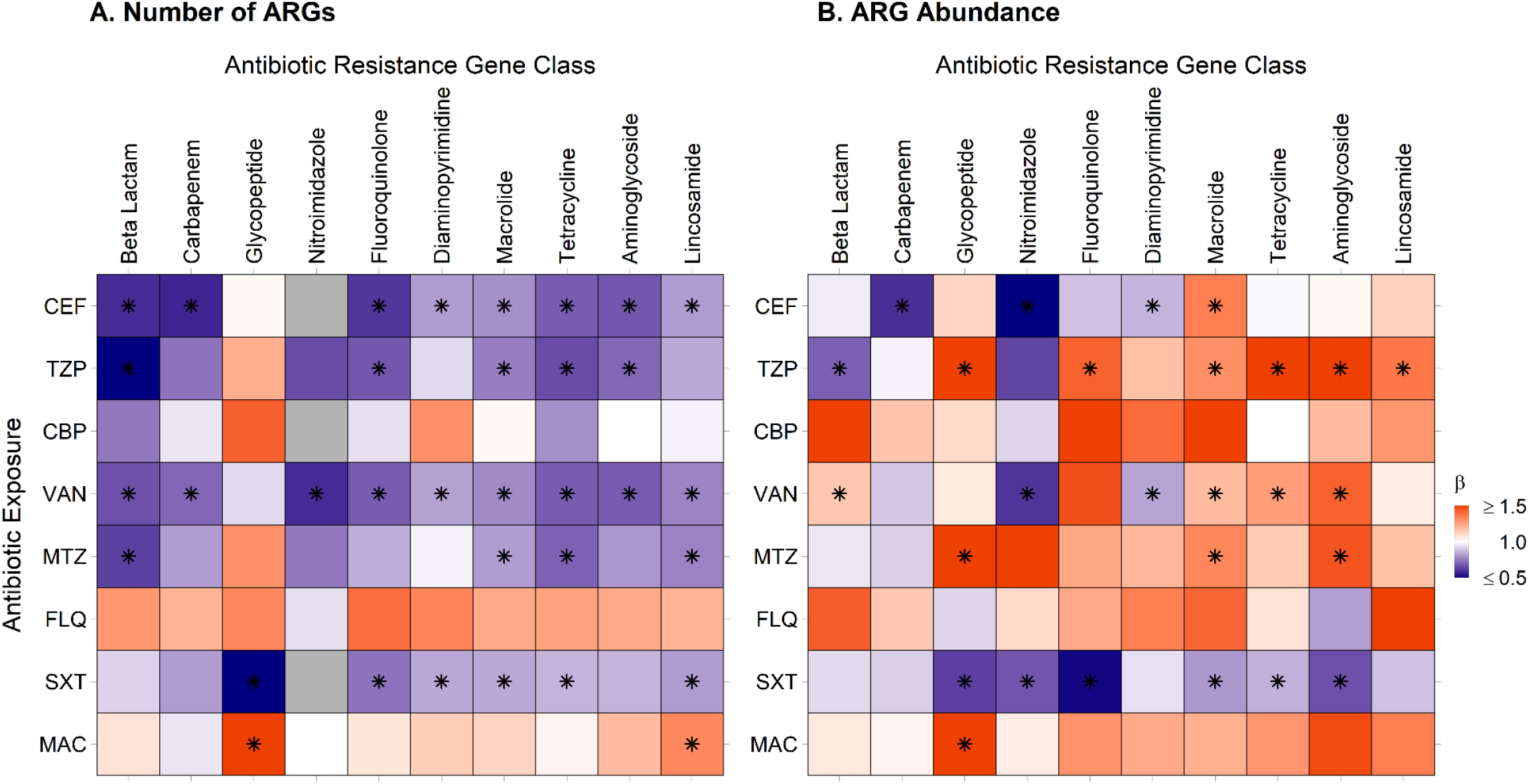
Heatmaps of clinically relevant antibiotic exposures’ effects on ARGs of the same and different antibiotic classes as the exposure. A. Antibiotic exposure effect on the number of ARGs for the exposure antibiotic class and others. B. Antibiotic exposure effect on the ARG abundance for the exposure antibiotic class and others. Beta indicates the effect size. ARGs, antibiotic resistance genes, FEP, cefepime; TZP, piperacillin-tazobactam; CBP, carbapenem; VAN, vancomycin; MTZ, metronidazole; FLQ, fluoroquinolone; SXT, trimethoprim-sulfamethoxazole; MAC, macrolide; Other, antibiotic exposure from tetracyclines, non-cefepime beta-lactams, clindamycin, and aminoglycosides. Tetracycline includes tetracycline and glycylcycline resistance genes; Beta-lactam includes cephalosporin, cephamycin, penam, and monobactam resistance genes; Lincosamide confers resistance to clindamycin; Carbapenem includes carbapenem and penem resistance genes; Glycopeptide confers resistance to vancomycin; Diaminopyrimidine confers resistance to trimethoprim; Nitromidazole confers resistance to metronidazole. Asterisks denote significant change. Gray boxes indicate there were not enough data to determine an effect.

## Discussion

Using shotgun metagenomics, we described the dynamic changes in the gut microbiome and resistome that occur among children undergoing HCT. We described the effects of antibiotics on both the composition of the gut microbiome and resistome, including the distinct effects of aerobic and anaerobic antibiotics on the relative abundances of bacterial species within the gut microbiome and on the number and abundance of specific ARG classes.

Compared to previous culture- and PCR-based methodologies, use of shotgun metagenomics for the characterization of the gut resistome is a novel way to study this complex, dynamic reservoir of ARGs.^21^ Considering ARGs collectively is particularly important because commensal bacteria, including those from the phyla Bacteroidetes and Firmicutes, likely harbor most of the ARGs in the healthy human gut and serve as a potential source for the horizontal transfer of ARGs to common enteric pathogens.^21,22^ Colonization by commensal, anaerobic bacteria with resistance genes may also explain how ARGs are detected even in the absence of antibiotic pressure.^23^ For instance, in our cohort, we detected a preponderance of ARGs that confer resistance to tetracyclines, macrolides, fluoroquinolones, and aminoglycosides despite the study population being infrequently exposed to these antibiotic classes. Knowledge of gut colonization by any bacteria with ARGs is especially important in the HCT population, as these patients are at high-risk of antibiotic-resistant bloodstream infections caused by gut-derived bacteria.

Surprisingly little is known about the dynamics of the gut resistome following HCT. A small cohort study of eight children undergoing HCT for high-risk acute leukemia found that the most commonly detected ARGs conferred resistance to tetracyclines, macrolides, beta-lactams, and aminoglycosides, as was seen in our cohort.^24^ They also concluded that the four children who developed acute GVHD during the study had a different resistome signature, with acquisition of several new ARGs after HCT, compared to children without GVHD.^24^ However, this study did not assess the effect of antibiotics on the gut resistome.^24^ Our findings suggest that antibiotics significantly impact both gut microbiome composition and the gut resistome throughout HCT. Due to the temporal similarities in the microbiome composition and resistome, the correlations between the microbiome and the resistome, and the similar loss of ARGs and species associated with antibiotic exposures, we concluded that changes to the resistome are largely impacted by changes in the microbiome composition. Specifically, antibiotic exposures lead to loss of bacterial species from the gut microbiome and this loss of species and their accompanying ARGs results in contraction of the gut resistome.

While both aerobic and anaerobic antibiotics were associated with a loss of microbiome and resistome richness, we found that aerobic and anaerobic antibiotics affected gut microbiome composition and the gut resistome differently. These findings are notable because anaerobic bacteria in the gut are primarily responsible for colonization resistance – the ability to prevent colonization by exogenous bacterial pathogens – through a variety of microbial and microbe-host interaction mechanisms.^25^ Thus, loss of anaerobic bacteria after anaerobic antibiotic exposures likely reduces colonization resistance, leading to expansion of existing antibiotic-resistant bacteria or the acquisition of exogenous antibiotic-resistant bacteria.^26^ Loss of colonization resistance with anaerobic antibiotics likely explains our findings of increased ARG acquisition, ARG abundance, and resistome instability despite the overall loss of ARGs associated with anaerobic antibiotics. When looking at specific anaerobic antibiotics, piperacillin-tazobactam and metronidazole were associated with lower relative abundances of *Bacteroides, Bifidobacterium, Blautia*, and *Clostridium* species, higher relative abundances of *Enterococcus* species, and an increased abundance of glycopeptide ARGs commonly found in enterococci. While we cannot directly assign ARGs to specific bacteria, these associations support the conclusion that exposure to these antibiotics results in loss of colonization resistance from anaerobes and colonization or expansion of *Enterococcus*. Taken together, our findings provide further evidence that anaerobic antibiotics should be used judiciously in this high-risk population.^9^

Our data also suggest that exposure to an antibiotic of one class can lead to acquisition of collateral antibiotic resistance to other antibiotic classes. In our cohort, vancomycin, piperacillin-tazobactam, and metronidazole exposures were associated with increases in the relative abundances of several other ARG classes, including fluoroquinolone, macrolide, tetracycline, and aminoglycoside ARGs. This phenomenon has been demonstrated in healthy adults, in which ciprofloxacin led to an increase in class D beta-lactamases and macrolide ARGs, and in premature neonates, in whom meropenem led to higher relative abundances of ARGs that confer resistance to fluoroquinolones, macrolides, tetracyclines, and trimethoprim.^27,28^ This expansion of collateral antibiotic resistance has likely also been observed clinically in the HCT population. In a meta-analysis of adults undergoing HCT, there was increased odds of a carbapenem-resistant *Klebsiella pneumoniae* infection not only with carbapenem exposure, but also with aminoglycoside, glycopeptide, and quinolone exposures.^29^ Similarly, infection with multidrug-resistant *Pseudomonas aeruginosa* was associated with vancomycin exposure in a separate meta-analysis of HCT recipients.^30^ Our data suggest that antibiotic pressure selects for organisms that are resistant to the exposure agent and also allows for the acquisition of new bacteria and expansion of existing bacteria with collateral ARGs of different antibiotic classes.

The effect of fluoroquinolone exposure on collateral antibiotic resistance is of particular interest, given that levofloxacin is the preferred agent for antibacterial prophylaxis after HCT.^31^ Fluoroquinolone use after HCT has been associated with a lower risk of fever and neutropenia and bloodstream infections.^31^ However, it has more recently been associated with increased risks of colonization and infection by carbapenem-resistant Enterobacteriaceae.^32^ In a murine model, ciprofloxacin exposure led to an increase in the abundance of beta-lactamase ARGs.^33^ We found increases in both the richness and abundance of several different ARG classes with fluoroquinolone exposure. However, none of these associations met statistical significance, likely an effect of the lack of statistical power with relatively few fluoroquinolone-exposed samples from our cohort. On the contrary, in a study of 49 children with acute lymphoblastic leukemia, levofloxacin prophylaxis was associated with an increase in the prevalence of fluoroquinolone resistance, while a rise in collateral antibiotic resistance from other ARG classes was not detected.^34^ As levofloxacin prophylaxis is becoming more common in pediatric centers, further research is needed to determine the effect of levofloxacin on the resistome.

Our study has several notable limitations. First, we are likely underestimating the effect of antibiotics on acquisition of ARGs given that antibiotic-induced changes in the gut microbiome are known to last weeks after cessation of antibiotic exposure.^35^ Second, as for many shotgun metagenomic sequencing analyses, the sequencing depth of our samples likely affected our ability to detect all ARGs present within a sample. To account for this, we included the sequencing depth in all models for our primary analyses. Unfortunately, analyses of associations between individual antibiotic exposures and ARG classes were not adjusted for sequencing depth because the adjusted models did not converge. Similarly, these models were not adjusted for time relative to HCT. Thus, while TMP-SMX exposure appears to reduce the acquisition of ARGs, this may reflect the fact that TMP-SMX was primarily administered prior to HCT and before the administration of broad-spectrum antibiotics for febrile neutropenia. Finally, with short-read sequencing data, we are unable to assign ARGs to specific bacterial species. Therefore, we can only make associations between gut microbiome composition and the gut resistome and cannot prove a direct link.

In conclusion, this study provides the largest, longitudinal evaluation of the pediatric gut resistome to date. Understanding the complex dynamics of the gut resistome during HCT and its response to antibiotic exposures can impact clinical decisions for antibiotic prophylaxis and treatment. This study provides further evidence for the importance of antimicrobial stewardship efforts within the HCT community. Antibiotic exposures in this high-risk population should be thoughtful in order to prevent further disruption of the gut microbiome and resistome and to avoid promoting antibiotic resistance.

## Data Availability

Data not included in the manuscript will be available by request to the corresponding authors. All metagenomic data will be uploaded to the Sequence Read Archive.

## Funding

Research reported in this publication was supported in part by the National Institute of Allergy and Infectious Diseases of the National Institutes of Health under Award Number UM1AI104681. The content is solely the responsibility of the authors and does not necessarily represent the official views of the National Institutes of Health. This work was additionally supported by the National Institute of Child Health and Human Development of the National Institutes of Health [T32HD094671 to S.M.H.], the National Human Genome Research Institute of the National Institutes of Health [T32 HG008955 to S.M.H.], the Translating Duke Health Ending Disease Where it Begins Working Group [to S.M.H.], the local efforts of the Duke Children’s Office of Development and its Children’s Miracle Network Hospitals fundraising corporate partnerships and programs [to S.M.H.], and a National Institutes of Health Career Development Award [K23-AI135090 to M.S.K.].

## Acknowledgements

We thank the Duke University School of Medicine for the use of the Microbiome Core Facility, which provided DNA extraction, and the Sequencing and Genomic Technologies Shared Resource, which provided library preparation and shotgun metagenomic sequencing. We especially thank all the children and families who participated in this study.

## Figure Legends

**Supplementary Table 1.**
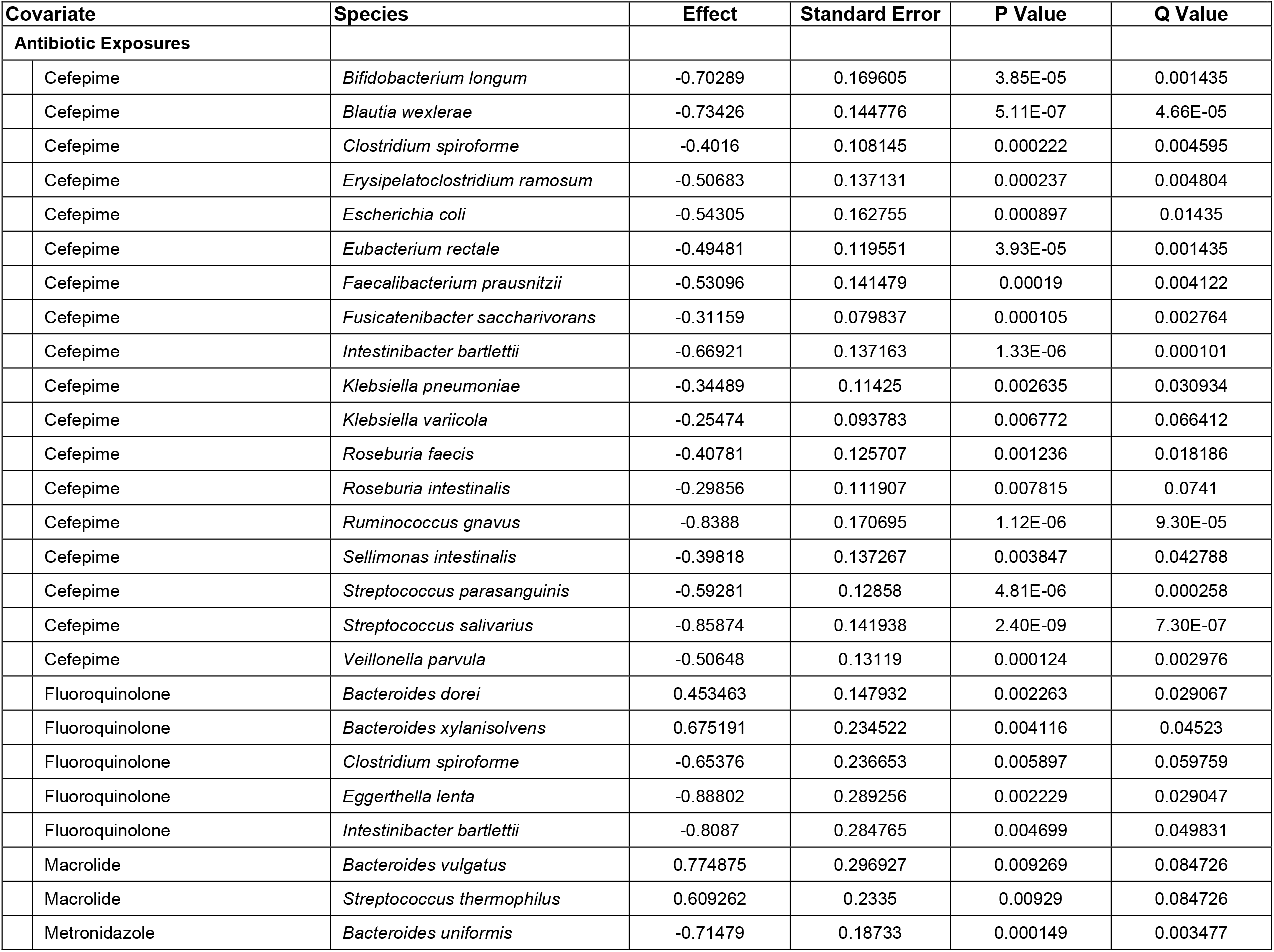
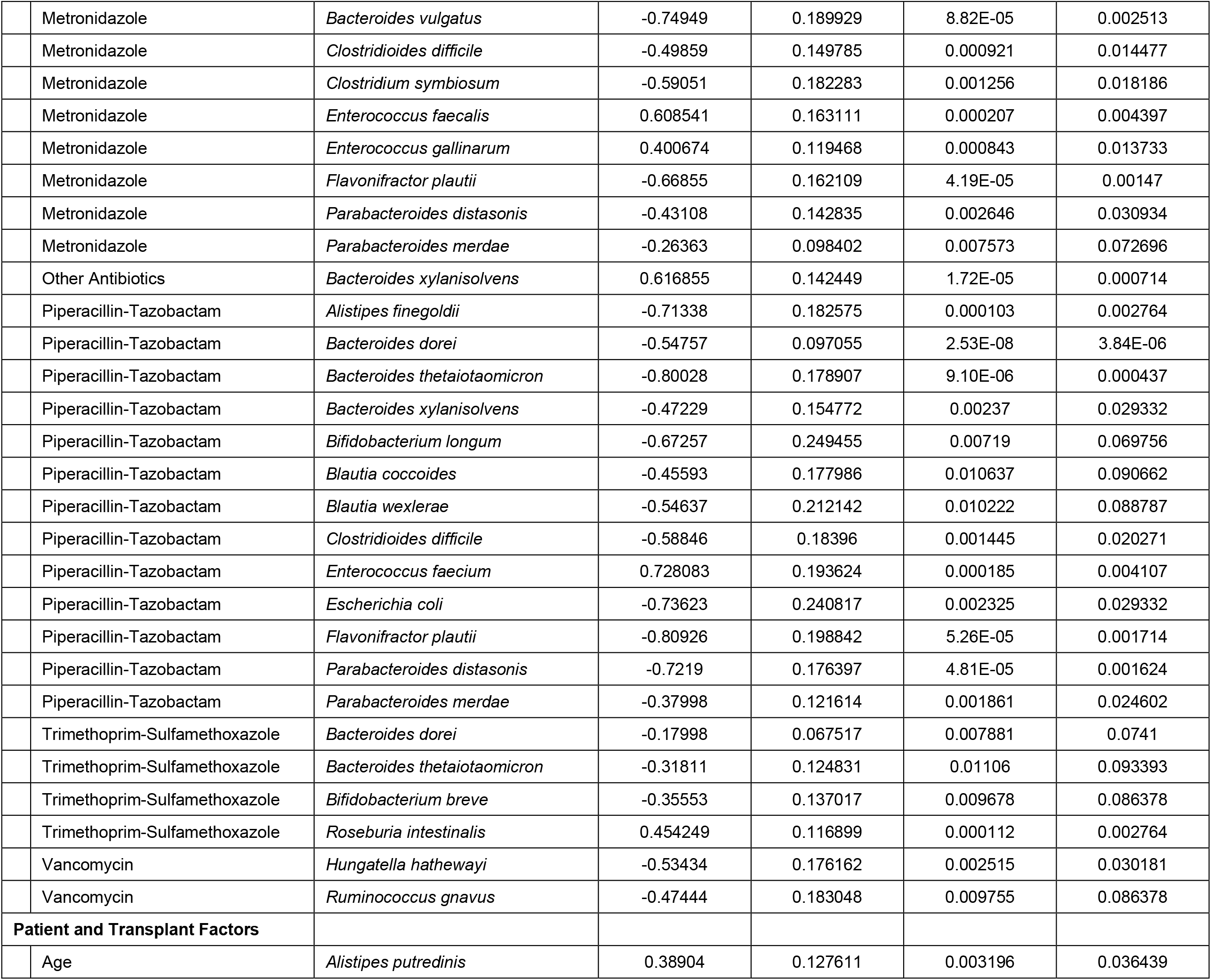
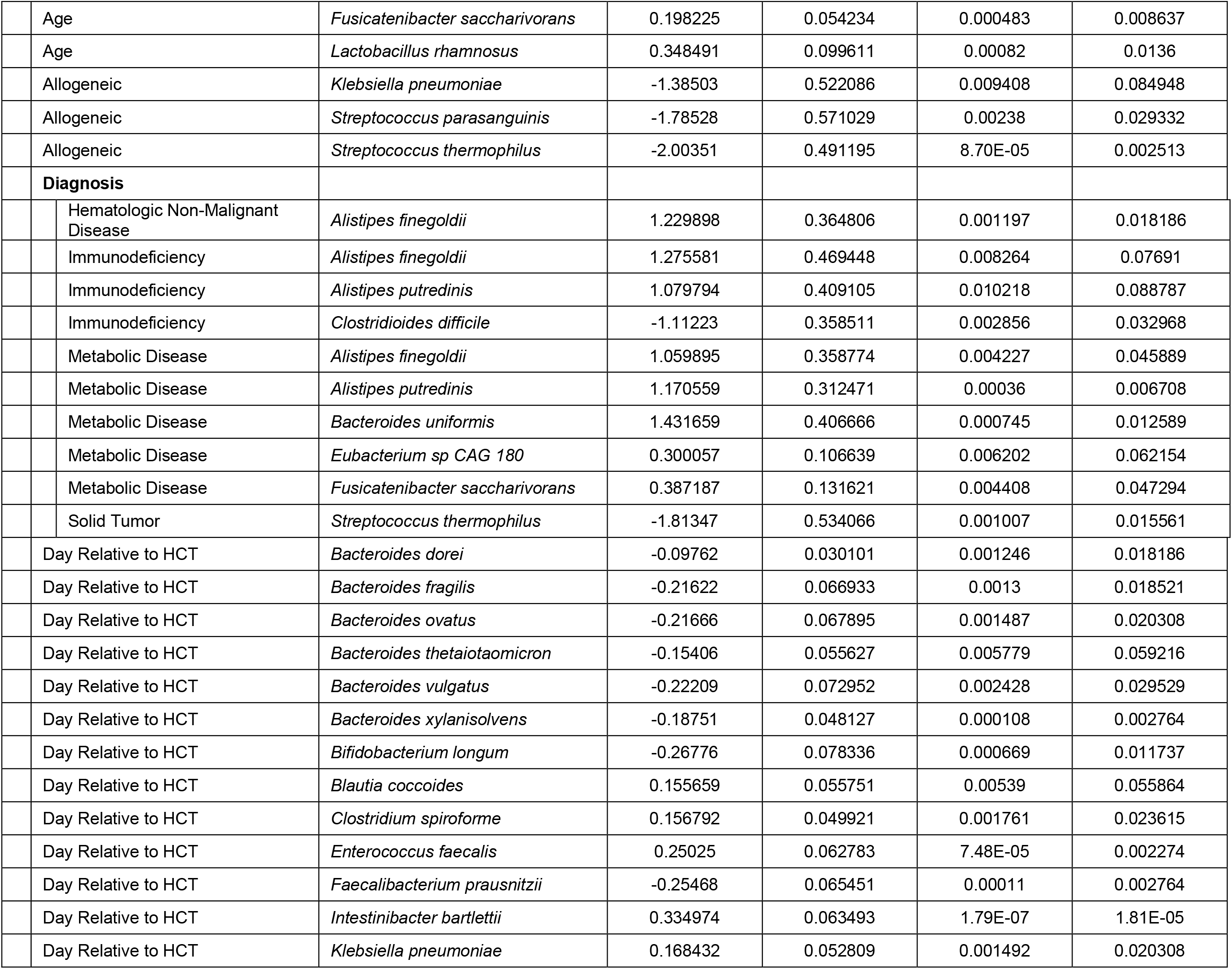
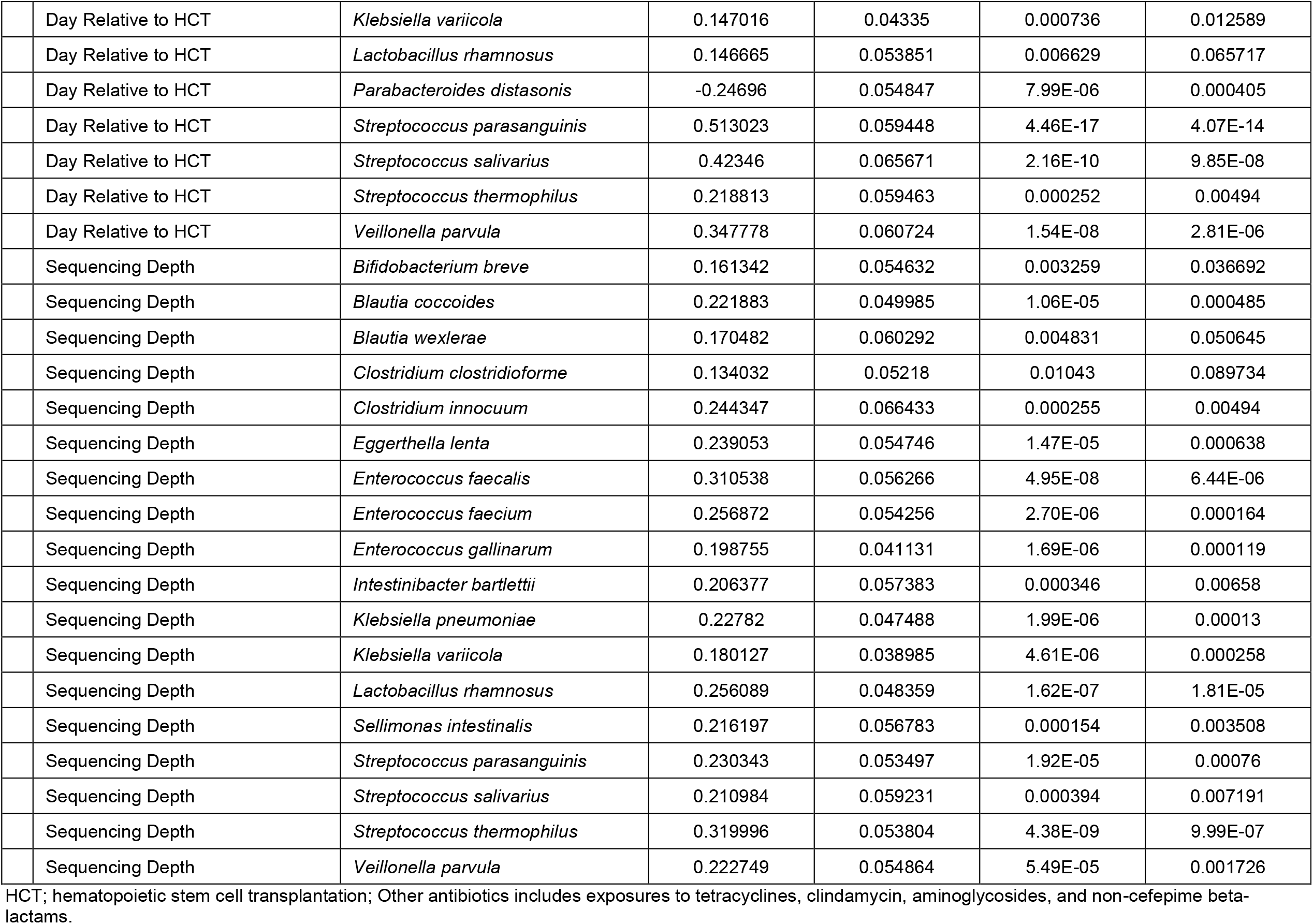
Significant associations of antibiotic exposures and clinical characteristics with abundances of bacterial species within the gut microbiome

**Supplementary Table 2.** Effect of clinical factors on measures of the resistome and microbiome.

| Clinical Factors | Resistome<br>$\beta$ (95% CI)<br>P value | | | | Microbiome<br>$\beta$ (95% CI)<br>P value | | |
| --- | --- | --- | --- | --- | --- | --- | --- |
|  | ARG Abundance | Number of ARGs | Number of New ARGs | Jaccard Distance ARGs | Number of Species | Number of New Species | Jaccard Distance Species |
| Aerobic Antibiotics | 1.13 (0.95, 1.35)<br>P=0.16 | 0.83 (0.76, 0.91)<br>P<0.0001 | 1.03 (0.88, 1.20)<br>P=0.74 | 0.02 (-0.03, 0.07)<br>P=0.47 | 0.72 (0.66, 0.79)<br>P<0.0001 | 0.95 (0.82, 1.11)<br>P=0.54 | 0.08 (0.04, 0.12)<br>P=0.0001 |
| Anaerobic Antibiotics | 1.95 (1.59, 2.38)<br>P=7.1e-11 | 0.84 (0.76, 0.93)<br>P=0.0007 | 1.29 (1.10, 1.52)<br>P=0.002 | 0.07 (0.03, 0.12)<br>P=0.003 | 0.68 (0.61, 0.76)<br>P<0.0001 | 1.19 (1.02, 1.39)<br>P=0.03 | 0.09 (0.05, 0.13)<br>P<0.0001 |
| Subject Age | 0.95 (0.92, 0.99)<br>P=0.02 | 1.00 (0.99, 1.02)<br>P=0.71 | 1.00 (0.97, 1.00)<br>P=0.10 | -0.01 (-0.01, -0.00)<br>P=0.03 | 1.03 (1.01, 1.05)<br>P=0.0004 | 1.00 (0.98, 1.02)<br>P=0.94 | -0.007 (-0.01, -0.00)<br>P=0.01 |
| Sex |  |  |  |  |  |  |  |
| Female | Reference |  |  |  | Reference |  |  |
| Male | 1.10 (0.74, 1.64)<br>P=0.63 | 1.07 (0.89, 1.29)<br>P=0.48 | 1.07 (0.91, 1.26)<br>P=0.40 | -0.01 (-0.07, 0.06)<br>P=0.84 | 0.99 (0.84, 1.17)<br>P=0.93 | 1.03 (0.87, 1.21)<br>P=0.76 | -0.02 (-0.08, 0.03)<br>P=0.38 |
| Diagnosis |  |  |  |  |  |  |  |
| Hematologic Malignancy | Reference |  |  |  | Reference |  |  |
| Congenital Immunodeficiency | 0.67 (0.33, 1.37)<br>P=0.27 | 1.21 (0.87, 1.70)<br>P=0.25 | 1.26 (0.94, 1.68)<br>P=0.12 | -0.02 (-0.14, 0.10)<br>P=0.72 | 1.51 (1.12, 2.05)<br>P=0.01 | 1.28 (0.95, 1.72)<br>P=0.10 | -0.08 (-0.17, 0.01)<br>P=0.11 |
| Non-malignant Heme | 1.15 (0.66, 2.00)<br>P=0.63 | 1.05 (0.81, 1.36)<br>P=0.73 | 0.97 (0.77, 1.23)<br>P=0.82 | -0.07 (-0.16, 0.02)<br>P=0.16 | 1.29 (1.02, 1.63)<br>P=0.04 | 1.06 (0.84, 1.34)<br>P=0.62 | -0.08 (-0.16, -0.01)<br>P=0.03 |
| Metabolic Disorder | 0.80 (0.46, 1.38)<br>P=0.42 | 0.97 (0.74, 1.26)<br>P=0.80 | 1.04 (0.82, 1.31)<br>P=0.77 | -0.05 (-0.14, 0.05)<br>P=0.36 | 1.49 (1.17, 1.88)<br>P=0.001 | 1.14 (0.90, 1.45)<br>P=0.28 | -0.08 (-0.15, -0.01)<br>P=0.05 |
| Solid Tumor | 0.41 (0.10, 1.64)<br>P=0.21 | 0.92 (0.48, 1.76)<br>P=0.80 | 0.75 (0.40, 1.42)<br>P=0.38 | -0.01 (-0.26, 0.23)<br>P=0.94 | 1.13 (0.63, 2.01)<br>P=0.68 | 0.72 (0.38, 1.35)<br>P=0.30 | -0.07 (-0.26, 0.12)<br>P=0.49 |
| Preparatory Regimen |  |  |  |  |  |  |  |
| Myeloablative | Reference |  |  |  | Reference |  |  |
| Non-myeloablative/RIC | 0.77 (0.26, 2.27)<br>P=0.64 | 0.97 (0.57, 1.63)<br>P=0.89 | 1.03 (0.60, 1.77)<br>P=0.92 | -0.03 (-0.22, 0.17)<br>P=0.79 | 1.13 (0.70, 1.81)<br>P=0.62 | 1.03 (0.60, 1.77)<br>P=0.90 | -0.06 (-0.21, 0.09)<br>P=0.45 |
| Type of HCT |  |  |  |  |  |  |  |
| Autologous | Reference |  |  |  | Reference |  |  |
| Allogeneic | 0.59 (0.17, 2.07)<br>P=0.41 | 0.75 (0.42, 1.35)<br>P=0.35 | 0.76 (0.42, 1.37)<br>P=0.36 | 0.05 (-0.18, 0.27)<br>P=0.68 | 0.61 (0.36, 1.03)<br>P=0.06 | 0.68 (0.38, 1.22)<br>P=0.20 | 0.03 (-0.14, 0.21)<br>P=0.74 |
| Day Relative to HCT | 1.003 (1.000, 1.006)<br>P=0.07 | 1.002 (1.001, 1.003)<br>P=0.006 | 1.008 (1.006, 1.010)<br>P<0.0001 | -0.001 (-0.002, -0.000)<br>P=0.03 | 1.001 (0.999, 1.002)<br>P=0.40 | 1.009 (1.007, 1.011)<br>P<0.0001 | -0.000 (-0.001, 0.000)<br>P=0.35 |
| Log of Sequencing Depth | 1.00 (0.91, 1.10)<br>P=0.98 | 1.33 (1.25, 1.40)<br>P<0.0001 | 1.36 (1.24, 1.50)<br>P<0.0001 | -0.03 (-0.05, -0.00)<br>P=0.04 | 1.35 (1.27, 1.42)<br>P<0.0001 | 1.38 (1.26, 1.52)<br>P<0.0001 | -0.04 (-0.06, -0.02)<br>P<0.0001 |

CI, confidence interval; ARG, antibiotic resistance gene; Non-malignant Heme, non-malignant hematologic disorder; RIC, reduced intensity conditioning; HCT, hematopoietic stem cell transplantation

**Supplementary Table 3.** Effect of antibiotics on the resistome.

| Antibiotic Exposure | Resistome<br>$\beta$ (95% CI)<br>P value | | | |
| --- | --- | --- | --- | --- |
|  | ARG Abundance | Number of ARGs | Number of New ARGs | Jaccard Distance ARGs |
| Cefepime | 1.35 (1.24, 1.47)<br>P<0.0001 | 0.78 (0.71, 0.86)<br>P<0.0001 | 1.29 (1.11, 1.50)<br>P=0.001 | 0.06 (0.02, 0.10)<br>P=0.01 |
| Vancomycin | 1.74 (1.62, 1.87)<br>P<0.0001 | 0.82 (0.74, 0.90)<br>P<0.0001 | 1.24 (1.06, 1.46)<br>P=0.01 | 0.06 (0.01, 0.10)<br>P=0.01 |
| Fluoroquinolone | 1.36 (0.88, 2.10)<br>P=0.17 | 1.13 (0.90, 1.42)<br>P=0.29 | 0.85 (0.60, 1.21)<br>P=0.37 | -0.10 (-0.21, 0.00)<br>P=0.07 |
| Aminoglycoside | 0.88 (0.50, 1.53)<br>P=0.64 | 0.88 (0.63, 1.23)<br>P=0.45 | 1.05 (0.63, 1.74)<br>P=0.86 | 0.11 (-0.03, 0.25)<br>P=0.11 |
| Macrolide | 1.16 (0.98, 1.38)<br>P=0.08 | 1.00 (0.82, 1.23)<br>P=0.97 | 1.14 (0.85, 1.54)<br>P=0.38 | 0.07 (-0.02, 0.16)<br>P=0.14 |
| TMP-SMX | 0.72 (0.65, 0.81)<br>P<0.0001 | 1.03 (0.92, 1.15)<br>P=0.62 | 0.84 (0.69, 1.03)<br>P=0.10 | 0.02 (-0.03, 0.08)<br>P=0.44 |
| Pip-Tazo | 3.19 (2.88, 3.53)<br>P<0.0001 | 0.78 (0.66, 0.91)<br>P=0.001 | 1.51 (1.20, 1.90)<br>P=0.0005 | 0.11 (0.03, 0.17)<br>P=0.004 |
| Carbapenem | 1.53 (1.27, 1.86)<br>P<0.0001 | 0.97 (0.74, 1.28)<br>P=0.85 | 1.15 (0.77, 1.72)<br>P=0.50 | 0.08 (-0.02, 0.19)<br>P=0.14 |
| Metronidazole | 1.84 (1.64, 2.06)<br>P<0.0001 | 0.85 (0.75, 0.97)<br>P=0.01 | 1.30 (1.08, 1.57)<br>P=0.007 | 0.09 (0.03, 0.14)<br>P=0.002 |
| Clindamycin | 1.39 (0.69, 2.78)<br>P=0.35 | 0.85 (0.57, 1.24)<br>P=0.39 | 1.01 (0.57, 1.78)<br>P=0.97 | 0.02 (-0.15, 0.17)<br>P=0.85 |

CI, confidence interval; ARG, antibiotic resistance gene; TMP-SMX, trimethoprim-sulfamethoxazole; Pip-Tazo, piperacillin-tazobactam

**Supplementary Table 4.** Effect of antibiotic exposure on the number of antibiotic resistance genes by gene class.

| Antibiotic Exposure | ARG Class<br>$\beta$ (95% CI)<br>Adjusted P value | | | | | | | | | |
| --- | --- | --- | --- | --- | --- | --- | --- | --- | --- | --- |
|  | Beta-Lactam | Carbapenem | Glycopeptide | Nitroimidazole | Fluoroquinolone | Diaminopyrimidine | Macrolide | Tetracycline | Aminoglycoside | Lincosamide |
| Cefepime | 0.63 (0.55, 0.73)<br>P<0.0001 | 0.62 (0.52, 0.75)<br>P<0.0001 | 1.02 (0.82, 1.28)<br>P=0.84 |  | 0.65 (0.56, 0.75)<br>P<0.0001 | 0.83 (0.74, 0.94)<br>P=0.005 | 0.81 (0.74, 0.89)<br>P<0.0001 | 0.72 (0.65, 0.79)<br>P<0.0001 | 0.71 (0.63, 0.80)<br>P<0.0001 | 0.83 (0.76, 0.92)<br>P=0.0003 |
| Pip-Tazo | 0.56 (0.44, 0.72)<br>P<0.0001 | 0.76 (0.56, 1.03)<br>P=0.11 | 1.23 (0.83, 1.80)<br>P=0.33 | 0.69 (0.44, 1.07)<br>P=0.12 | 0.71 (0.56, 0.89)<br>P=0.01 | 0.93 (0.76, 1.15)<br>P=0.53 | 0.78 (0.65, 0.92)<br>P=0.01 | 0.69 (0.58, 0.81)<br>P<0.0001 | 0.74 (0.60, 0.91)<br>P=0.01 | 0.85 (0.71, 1.00)<br>P=0.08 |
| Carbapenem | 0.77 (0.48, 1.22)<br>P=0.60 | 0.95 (0.55, 1.64)<br>P=1.00 | 1.43 (0.78, 2.64)<br>P=0.60 |  | 0.94 (0.63, 1.40)<br>P=1.00 | 1.31 (0.97, 1.77)<br>P=0.37 | 1.02 (0.77, 1.35)<br>P=1.00 | 0.81 (0.60, 1.09)<br>P=0.49 | 0.99 (0.69, 1.42)<br>P=1.00 | 0.97 (0.73, 1.27)<br>P=1.00 |
| Vancomycin | 0.70 (0.60, 0.82)<br>P<0.0001 | 0.74 (0.61, 0.90)<br>P=0.005 | 0.93 (0.74, 1.18)<br>P=0.57 | 0.63 (0.47, 0.85)<br>P=0.005 | 0.72 (0.61, 0.84)<br>P<0.0001 | 0.84 (0.74, 0.96)<br>P=0.02 | 0.80 (0.72, 0.89)<br>P<0.0001 | 0.72 (0.65, 0.80)<br>P<0.0001 | 0.72 (0.63, 0.82)<br>P<0.0001 | 0.79 (0.71, 0.88)<br>P<0.0001 |
| Metronidazole | 0.67 (0.53, 0.84)<br>P=0.003 | 0.83 (0.64, 1.08)<br>P=0.20 | 1.31 (0.97, 1.78)<br>P=0.16 | 0.77 (0.53, 1.11)<br>P=0.20 | 0.86 (0.70, 1.05)<br>P=0.20 | 0.97 (0.82, 1.14)<br>P=0.69 | 0.83 (0.72, 0.96)<br>P=0.03 | 0.73 (0.63, 0.85)<br>P=0.0007 | 0.82 (0.69, 0.98)<br>P=0.07 | 0.79 (0.69, 0.91)<br>P=0.005 |
| Fluoroquinolone | 1.29 (0.90, 1.86)<br>P=0.28 | 1.21 (0.77, 1.91)<br>P=0.59 | 1.33 (0.63, 2.82)<br>P=0.60 | 0.94 (0.51, 1.75)<br>P=0.85 | 1.40 (0.97, 2.01)<br>P=0.26 | 1.34 (0.98, 1.83)<br>P=0.26 | 1.24 (0.96, 1.59)<br>P=0.27 | 1.26 (0.99, 1.60)<br>P=0.26 | 1.24 (0.91, 1.68)<br>P=0.28 | 1.21 (0.95, 1.54)<br>P=0.28 |
| TMP-SMX | 0.92 (0.78, 1.08)<br>P=0.33 | 0.83 (0.68, 1.02)<br>P=0.10 | 0.56 (0.42, 0.75)<br>P=0.0005 |  | 0.76 (0.64, 0.90)<br>P=0.006 | 0.85 (0.74, 0.98)<br>P=0.04 | 0.85 (0.76, 0.96)<br>P=0.02 | 0.87 (0.78, 0.97)<br>P=0.02 | 0.87 (0.76, 1.01)<br>P=0.08 | 0.83 (0.75, 0.93)<br>P=0.004 |
| Macrolide | 1.08 (0.77, 1.51)<br>P=0.89 | 0.95 (0.64, 1.43)<br>P=0.89 | 2.07 (1.35, 3.16)<br>P=0.01 | 1.00 (0.58, 1.74)<br>P=0.99 | 1.07 (0.77, 1.48)<br>P=0.89 | 1.14 (0.87, 1.48)<br>P=0.57 | 1.12 (0.90, 1.39)<br>P=0.57 | 1.03 (0.83, 1.29)<br>P=0.89 | 1.19 (0.91, 1.55)<br>P=0.44 | 1.33 (0.10, 1.61)<br>P=0.02 |
| Other° | 1.07 (0.85, 1.35)<br>P=0.80 | 1.27 (0.96, 1.68)<br>P=0.48 | 1.16 (0.77, 1.75)<br>P=0.80 | 1.09 (0.74, 1.59)<br>P=0.80 | 1.03 (0.81, 1.30)<br>P=0.88 | 1.11 (0.91, 1.34)<br>P=0.67 | 1.02 (0.86, 1.20)<br>P=0.88 | 0.99 (0.85, 1.17)<br>P=0.95 | 1.15 (0.94, 1.40)<br>P=0.48 | 0.94 (0.70, 1.11)<br>P=0.80 |

°Other includes clindamycin, tetracycline, non-cefepime beta-lactams, and aminoglycoside exposures.
ARG, antibiotic resistance gene; CI, confidence interval; Pip-Tazo, piperacillin-tazobactam; TMP-SMX, trimethoprim-sulfamethoxazole

**Supplementary Table 5.** Effect of antibiotic exposure on the abundance of antibiotic resistance genes by gene class.

| Antibiotic Exposure | ARG Class<br>β (95% CI)<br>Adjusted P value |  |  |  |  |  |  |  |  |  |
| --- | --- | --- | --- | --- | --- | --- | --- | --- | --- | --- |
|  | Beta-Lactam | Carbapenem | Glycopeptide | Nitroimidazole | Fluoroquinolone | Diaminopyrimidine | Macrolide | Tetracycline | Aminoglycoside | Lincosamide |
| Cefepime | 0.91 (0.77, 1.07)<br>P=0.33 | 0.58 (0.47, 0.73)<br>P<0.0001 | 1.13 (0.88, 1.46)<br>P=0.38 | 0.44 (0.34, 0.58)<br>P<0.0001 | 0.84 (0.72, 0.99)<br>P=0.06 | 0.82 (0.70, 0.97)<br>P=0.04 | 1.36 (1.11, 1.66)<br>P=0.005 | 0.93 (0.82, 1.05)<br>P=0.33 | 1.04 (0.89, 1.21)<br>P=0.72 | 1.14 (0.99, 1.31)<br>P=0.79 |
| Pip-Tazo | 0.67 (0.51, 0.87)<br>P=0.01 | 0.92 (0.66, 1.30)<br>P=0.65 | 11.80 (8.78, 15.85)<br>P<0.0001 | 0.62 (0.40, 0.97)<br>P=0.06 | 1.43 (1.16, 1.77)<br>P=0.003 | 1.19 (0.88, 1.60)<br>P=0.29 | 1.32 (1.08, 1.62)<br>P=0.01 | 3.37 (2.84, 4.01)<br>P<0.0001 | 2.70 (2.25, 3.25)<br>P<0.0001 | 1.38 (1.08, 1.76)<br>P=0.02 |
| Carbapenem | 1.57 (1.04, 2.36)<br>P=0.20 | 1.18 (0.63, 2.23)<br>P=1.00 | 1.12 (0.54, 2.32)<br>P=1.00 | 0.87 (0.38, 2.00)<br>P=1.00 | 1.69 (1.16, 2.46)<br>P=0.13 | 1.41 (0.93, 2.13)<br>P=0.40 | 1.50 (1.08, 2.09)<br>P=0.16 | 1.02 (0.70, 1.48)<br>P=1.00 | 1.20 (0.83, 1.74)<br>P=0.66 | 1.30 (0.90, 1.87)<br>P=0.49 |
| Vancomycin | 1.17 (1.01, 1.36)<br>P=0.047 | 0.85 (0.67, 1.07)<br>P=0.20 | 1.08 (0.82, 1.43)<br>P=0.57 | 0.59 (0.44, 0.80)<br>P=0.001 | 1.47 (1.02, 2.13)<br>P=0.05 | 0.79 (0.66, 0.96)<br>P=0.02 | 1.20 (1.05, 1.37)<br>P=0.01 | 1.28 (1.13, 1.45)<br>P=0.0002 | 1.43 (1.24, 1.65)<br>P<0.0001 | 1.07 (0.92, 1.25)<br>P=0.41 |
| Metronidazole | 0.90 (0.72, 1.12)<br>P=0.36 | 0.86 (0.63, 1.17)<br>P=0.36 | 1.67 (1.16, 2.40)<br>P=0.02 | 1.68 (0.76, 3.72)<br>P=0.24 | 1.26 (0.98, 1.60)<br>P=0.15 | 1.21 (0.96, 1.52)<br>P=0.17 | 1.34 (1.11, 1.62)<br>P=0.008 | 1.16 (0.96, 1.39)<br>P=0.018 | 1.46 (1.19, 1.81)<br>P=0.003 | 1.19 (0.97, 1.46)<br>P=0.16 |
| Fluoroquinolone | 1.44 (0.96, 2.16)<br>P=0.26 | 1.17 (0.64, 2.14)<br>P=0.75 | 0.87 (0.37, 2.02)<br>P=0.79 | 1.12 (0.56, 2.24)<br>P=0.79 | 1.25 (0.87, 1.81)<br>P=0.35 | 1.36 (0.90, 2.07)<br>P=0.28 | 1.42 (1.02, 1.98)<br>P=0.26 | 1.10 (0.69, 1.76)<br>P=0.79 | 0.78 (0.57, 1.08)<br>P=0.28 | 1.53 (1.08, 2.18)<br>P=0.26 |
| TMP-SMX | 0.88 (0.72, 1.08)<br>P=0.24 | 0.86 (0.67, 1.10)<br>P=0.24 | 0.61 (0.44, 0.86)<br>P=0.01 | 0.65 (0.48, 0.89)<br>P=0.02 | 0.51 (0.42, 0.62)<br>P<0.0001 | 0.89 (0.74, 1.07)<br>P=0.24 | 0.77 (0.66, 0.90)<br>P=0.004 | 0.81 (0.70, 0.95)<br>P=0.002 | 0.64 (0.53, 0.77)<br>P<0.0001 | 0.84 (0.71, 0.99)<br>P=0.06 |
| Macrolide | 1.08 (0.77, 1.51)<br>P=0.89 | 1.05 (0.65, 1.68)<br>P=0.89 | 3.50 (2.11, 5.79)<br>P<0.0001 | 1.07 (0.61, 1.86)<br>P=0.89 | 1.31 (0.94, 1.84)<br>P=0.31 | 1.25 (0.87, 1.80)<br>P=0.44 | 1.23 (0.93, 1.64)<br>P=0.38 | 1.30 (0.98, 1.72)<br>P=0.22 | 1.48 (1.08, 2.04)<br>P=0.08 | 1.36 (1.00, 1.83)<br>P=0.20 |
| Other° | 0.58 (0.46, 0.75)<br>P=0.0004 | 1.43 (1.01, 2.03)<br>P=0.43 | 1.09 (0.71, 1.70)<br>P=0.80 | 1.11 (0.72, 1.70)<br>P=0.80 | 1.22 (0.69, 2.17)<br>P=0.80 | 1.24 (0.96, 1.61)<br>P=0.48 | 1.17 (0.93, 1.47)<br>P=0.48 | 1.06 (0.85, 1.31)<br>P=0.80 | 0.82 (0.60, 1.11)<br>P=0.48 | 1.18 (0.94, 1.49)<br>P=0.48 |

°Other includes clindamycin, tetracycline, non-cefepime beta-lactams, and aminoglycoside exposures.
ARG, antibiotic resistance gene; CI, confidence interval; Pip-Tazo, piperacillin-tazobactam; TMP-SMX, trimethoprim-sulfamethoxazole

## Notes

### Competing Interest Statement

The authors have declared no competing interest.

### Author Declarations

We obtained informed consent from participants legal guardians prior to enrollment, and the study protocol was approved by the Duke University Health System Institutional Review Board.

